# Effects of a 12-Week Mat Pilates Program on Dietary Acid Load, Mental Well-Being, and Physical Fitness in Female Athletes

**DOI:** 10.1101/2025.06.05.25329092

**Authors:** Emrah Atay, Hilal Ertürk Yaşar, Didem Gülçin Kaya

**Affiliations:** Department of Physical Education and Sports, Faculty of Sport Science, Burdur Mehmet Akif Ersoy University, Burdur, Turkey; Department of Physical Education and Sports, Faculty of Sport Science, Afyon Kocatepe University, Afyonkarahisar, Turkey; Department of Physical Education and Sports, Faculty of Sport Science, Gazi University, Ankara, Turkey

**Keywords:** Acid-base balance, PRAL, Pilates Exercise, Mental Well-Being, Physical activity

## Abstract

This study aimed to evaluate the effects of a 12-week Pilates program on dietary acid load (PRAL), body composition, physical performance, and mental well-being in female athletes, focusing on the relationship between dietary acid load and physiological and psychological outcomes. Seventeen female athletes participated in Pilates sessions three times per week for 12 weeks. Dietary intake was assessed using 3-day food diaries at baseline and post-intervention to calculate PRAL values. Hydration was monitored through urine pH and specific gravity. Anthropometric measures included BMI, body fat percentage, waist circumference, fat mass, lean mass, and grip strength. Mental well-being was assessed with the Warwick-Edinburgh Mental Well-Being Scale, and physical activity perception was evaluated using the Cognitive Behavioral Physical Activity Questionnaire. Measurements were taken before and after the intervention. After the program, significant improvements were observed in grip strength, mental well-being, and physical activity perception. Small increases in BMI, body fat percentage, and waist circumference were noted. A significant correlation was found between PRAL values and both physical and psychological variables. Pilates positively affects mental well-being, physical activity engagement, and interacts with diet-related acid-base balance and body composition. Pilates may improve both physical and mental health in active female populations.

## 1. Introductıon

In today’s world, individuals who recognize the negative impacts of a sedentary lifestyle are increasingly turning to physical activity. Particularly in the context of the benefits brought by modern advancements, current living conditions—while making life more convenient—also contribute to physical limitations, serving as a catalyst for this shift towards more active lifestyles (1). Physical inactivity, driven by increased vehicle use and poor eating habits, coupled with the growing preference for ready-to-eat foods as a means of saving time, exemplifies the aforementioned living conditions. In this context, Pilates has become one of the most widely preferred exercises for improving posture disorders (2) for managing various diseases and pain that may arise in the future (3). Due to its growing popularity and widespread preference, the use of Pilates is increasingly being incorporated into sports sciences and physiotherapy.

Developed by Joseph Hubertus Pilates in the early 20th century, Pilates is an exercise method designed to strengthen muscles, enhance flexibility, and promote overall health by maintaining the balance between the mind and body (4,5). In addition to the breathing technique, the Pilates exercise also targets mental focus (6). Pilates is often referred to as “Contrology” or “control science” because it emphasizes the development of both mental control over the muscles and physical improvement, incorporating the principles of strength, balance, flexibility, posture, and breathing (7,8). Initially used during World War I to rehabilitate individuals held in prison camps and those injured in battle, this exercise method later gained popularity among ballerinas, ballet dancers, and dance instructors due to its therapeutic effects (4,9). Over time, this exercise method has been passed down through generations of instructors and continues to be practiced today. Pilates classes are now offered as part of elective and specialized courses in the field of sports sciences to encourage physical activity among sedentary individuals, promote public health, and improve the general health of all age groups, from infants to adults. Additionally, experts in the field are trained to provide these classes.

Physical activity refers to the energy expenditure resulting from the movement of the body through skeletal muscles (10). Activities such as climbing stairs and walking, which can be performed with or without a plan at any time and in any location, are examples of physical activity (11). Therefore, both planned and unplanned Pilates exercises are considered forms of physical activity and play an essential role in minimizing the risk of various diseases. Vural et al. (2010) emphasized that physical activity levels and nutritional status are crucial factors in enhancing individuals’ quality of life and improving their overall health (12). Similarly, the World Health Organization (2004) has highlighted that physical activity and a healthy diet are protective factors against cardiovascular disease, diabetes, and depression (13). While many individuals focus on physical activity, there is also increasing attention to healthy eating. However, a review of the literature suggests that physical activity alone may not be sufficient; it is equally important to pay attention to maintaining a healthy diet.

Nutrition refers to the optimal intake and utilization of macro and micronutrients required by the body to support growth, development, and maintain overall health (14). For healthy eating behaviors to occur, an individual must consume all necessary nutrients in an adequate and balanced manner (15). In sports nutrition, sufficient and balanced nutrition is personalized, taking into account factors such as age, gender, habits, environmental influences, physical activity, and energy expenditure of the athlete (16). In this context, the acid-base balance in body fluids, which is influenced by nutrition, is also considered (17). This is where the concept of dietary acid load comes into play, depending on dietary choices. Protein-rich foods, such as meat, fish, milk, dairy products, eggs, whole grains, and processed foods, are also rich in phosphorus and sulfur, which tend to increase the acidic load. On the other hand, diets rich in calcium, magnesium, and potassium contribute to an increased alkaline load(18).

In this regard, the formula commonly used in the literature to calculate the acid-base balance created by foods in the body and to determine dietary acid load is the Renal Acid Load (PRAL) value (19,20). Athletes are typically advised to consume protein and carbohydrates to meet their energy requirements for exercise, which results in higher PRAL values (21).

Thus, it is predicted that student-athletes who pilate regularly and know about healthy eating tend to have high PRAL values. In addition, it is thought that their mental well-being level is high because they are aware of their abilities, develop physically and mentally, and increase their quality of life. Mental well-being is a condition that allows individuals to realize their talents, contribute to society, and cope with difficulties and stress (13). In this context, the mental well-being and physical activity levels of athlete students, who will educate the next generation, are crucial and are influenced by the type of exercise they engage in. Specifically, it is essential to examine the development of athlete students through experimental practice to increase the number of individuals participating in physical activity, encourage healthy eating habits, and guide societies toward proper physical activity engagement, thus contributing to societal development. Therefore, this study aimed to assess the dietary acid loads of students from the Faculty of Sports Sciences who regularly practice Pilates, and to explore the impact of Pilates exercises on mental well-being and physical activity levels.

## 2. Method

### 2.1. Research Design

This study employed a descriptive cross-sectional design and was conducted in accordance with the Declaration of Helsinki. Initially, all students enrolled in the Pilates training program at the Faculty of Sports Sciences were considered for participation. However, four individuals were excluded due to not meeting the inclusion criteria or having specific health conditions. The final sample comprised 17 female athletes.

Prior to the intervention, all assessments outlined in the data collection instruments were conducted by a registered dietitian and a certified trainer. The study was implemented in two phases:

Phase 1: Participants received detailed information about the study, and both verbal and written informed consent were obtained to confirm voluntary participation. Baseline anthropometric and body composition measurements were recorded. Participants then engaged in supervised mat-based Pilates sessions three times per week for 12 weeks. On non-training days, they were instructed to avoid strenuous physical activity. The Pilates sessions were conducted under expert supervision in accordance with the standards set by the Pilates Committee of the Turkish Gymnastics Federation. Participant recruitment began on 2 January 2024 and continued until 25 March 2024, covering the full 12-week intervention period. All participants were aged 18 or older and provided both verbal and written informed consent. Prior to the study, group meetings were held with all athletes to explain the procedures in detail, and those who voluntarily agreed to participate signed a written consent form. Verbal consent was obtained in these sessions and was recorded in the study logbook. No minors were included, and the ethics committee required and approved the consent process.

Phase 2: Post-intervention anthropometric and body composition assessments were repeated for participants who completed the program. Additionally, changes in physical and mental well-being were evaluated at the end of the 12-week intervention.

Ethical approval for the study was granted by the Scientific Research and Publication Ethics Committee of a university affiliated with one of the authors (Non-Interventional Clinical Research Ethics Committee Decision; Meeting Date: 03.12.2024, Meeting No: 2024/12, Decision No: GO 2024/639).

### 2.2. Research Group

The study sample consisted of 17 female athletes with a mean age of 20.71 ± 1.86 years. The average height of the participants was 1.66 ± 0.07 meters. Inclusion criteria were: no diagnosed health issues, signing of the informed consent form, and no use of drugs or anabolic steroids within the past six months.

### 2.3. Data Collection Tools

#### Determination of Dietary Acid Load

Participants were instructed to record a 3-day food diary, including at least one day of Pilates training. All dietary records were completed under the supervision of a registered dietitian to ensure accuracy. The dietary intake data were analyzed using the BeBiS Nutrition Software (version 9.0), a validated tool for nutritional assessment.

The Potential Renal Acid Load (PRAL) was calculated using the following equation:

PRAL (mEq/100 g) = 0.49 × Protein (g) + 0.037 × Phosphorus (mg) − 0.021 × Potassium (mg) − 0.026 × Magnesium (mg) − 0.013 × Calcium (mg)

Based on individual PRAL scores, participants were categorized into two groups:

- **Low PRAL group:** PRAL < 15 mEq/day
- **High PRAL group:** PRAL ≥ 15 mEq/day (22).

#### Urine Assessment

Following completion of the 3-day dietary records, morning urine samples were collected after an overnight fast. Urine pH values were interpreted as follows: ≤6.5 as acidic, 7.0 as neutral, and ≥7.5 as alkaline. These values were used to assess participants’ acid-base balance.

Hydration status was assessed using urine specific gravity (USG). A USG ≤1.020 indicated euhydration, values between 1.021 and 1.030 indicated hypohydration, and values >1.030 were classified as severe hypohydration. (23,24).

#### Anthropometric Characteristics and Body Composition Evaluation

Body weight, body fat mass, lean body mass, body fat percentage, body water percentage, and bone mass (kg) were assessed using a Tanita MC-980 multi-frequency bioelectrical impedance analyzer. Measurements were taken with participants wearing minimal clothing and without shoes or socks.

Height was measured using a Leicester portable stadiometer. Waist circumference was determined with a non-elastic tape measure at the midpoint between the lowest rib and the iliac crest, with participants wearing light clothing.

Hip circumference was measured at the widest and most prominent point of the buttocks. The waist-to-hip ratio (WHR) was then calculated using the formula:

WHR = Waist Circumference (cm) / Hip Circumference (cm)

#### Lower Body Flexibility

Flexibility was measured using the sit-and-reach (SR) test. Participants reached forward as far as possible without bending their knees or torso. The test was repeated twice, and the best result was recorded.

#### Handgrip Strength

Hand and forearm muscle strength, including claw strength, was assessed using a standard dynamometer. Measurements followed the American Association of Hand Therapists (AHT-AETD) protocol: participants were seated with the shoulder adducted and in neutral rotation, elbow flexed at 90°, forearm in a neutral supported position, and wrist in neutral. Three measurements were taken with one-minute rest intervals; the average value was recorded. (25).

#### Evaluation of Physical and Mental Changes in Individuals After Exercise

- **The Cognitive Behavioral Physical Activity Questionnaire:** Developed by Schembre et al. (2015) and adapted to Turkish by Eskiler et al. (2016), this five-point Likert-type scale consists of 15 items across three sub-dimensions: outcome expectation, self-regulation, and personal barriers.(26).
- **Warwick-Edinburgh Mental Well-Being Scale Short Form:** Developed by Tennant et al. (2007) and adapted to Turkish by Demirtaş and Baytemir (2019). This five-point Likert-type scale consists of 7 items. Participants responded based on their experiences over the past two weeks (27,28).

### 2.4. Statistical analysis

Statistical analyses were conducted using SPSS Statistics software (version 27, IBM Corp., Armonk, NY, USA). Descriptive statistics, including means and standard deviations (SD), were used to summarize participants’ demographic characteristics. The Shapiro–Wilk test was employed to assess the normality of the data distribution.

For variables not normally distributed, the Wilcoxon signed-rank test was applied. Relationships between categorical variables were examined using the Chi-square test of independence. All results are reported as means ± standard error (SE), unless otherwise specified. A p-value of ≤0.05 was considered statistically significant.

## 3. Results

The study included 17 female athletes with an average age of 20.71 ± 1.86 years. The mean BMI of the participants was 20.98 ± 2.76. Demographic information of the participants is presented in Table 1.

**Table 1.**
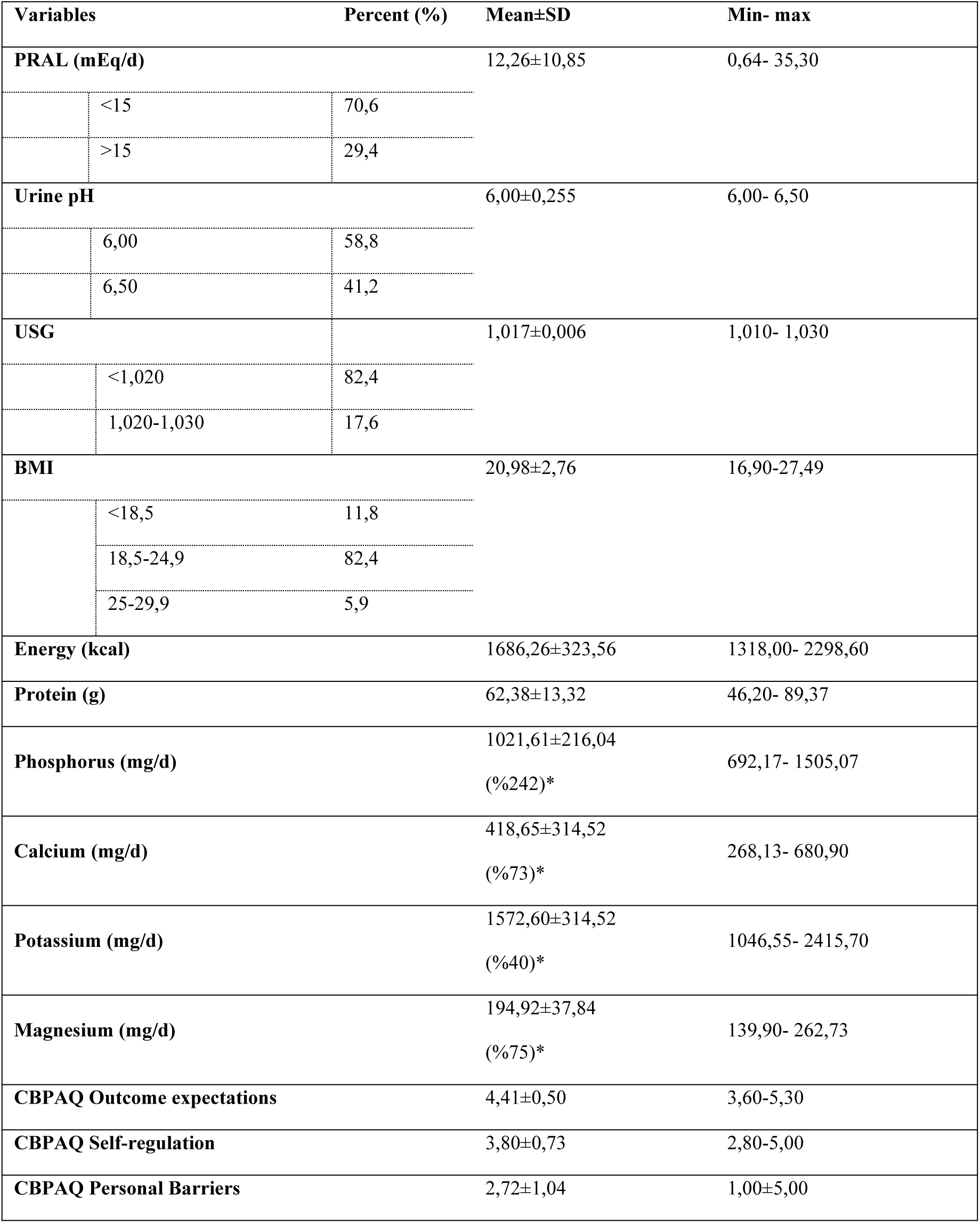
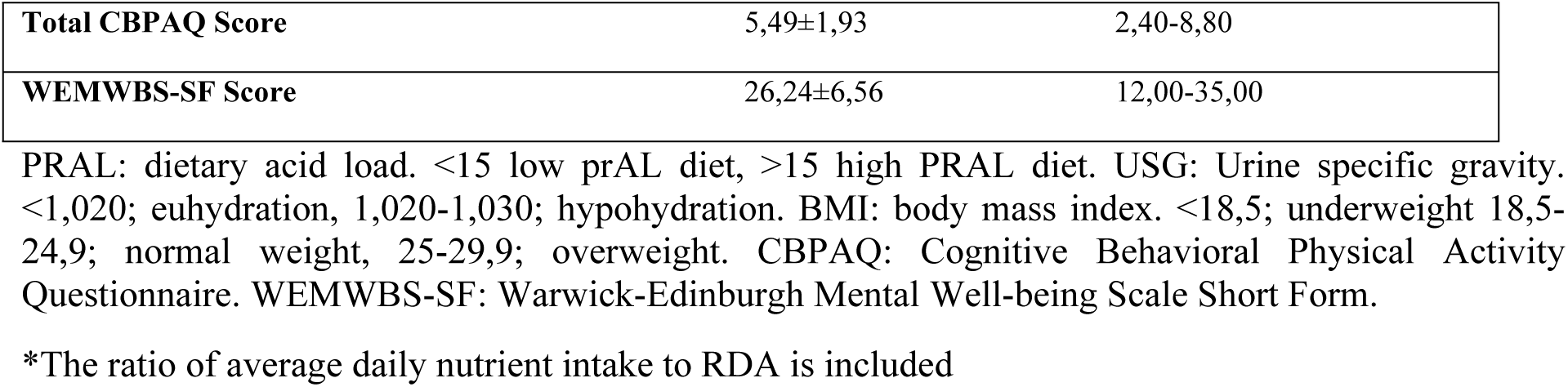
Descriptive information of participants.

| Variables | | Percent (%) | Mean±SD | Min- max |
| --- | --- | --- | --- | --- |
| **PRAL (mEq/d)** | | | 12,26±10,85 | 0,64- 35,30 |
|  | <15 | 70,6 |  |  |
|  | >15 | 29,4 |  |  |
| **Urine pH** | | | 6,00±0,255 | 6,00- 6,50 |
|  | 6,00 | 58,8 |  |  |
|  | 6,50 | 41,2 |  |  |
| **USG** | | | 1,017±0,006 | 1,010- 1,030 |
|  | <1,020 | 82,4 |  |  |
|  | 1,020-1,030 | 17,6 |  |  |
| **BMI** | | | 20,98±2,76 | 16,90-27,49 |
|  | <18,5 | 11,8 |  |  |
|  | 18,5-24,9 | 82,4 |  |  |
|  | 25-29,9 | 5,9 |  |  |
| **Energy (kcal)** | | | 1686,26±323,56 | 1318,00- 2298,60 |
| **Protein (g)** | | | 62,38±13,32 | 46,20- 89,37 |
| **Phosphorus (mg/d)** | | | 1021,61±216,04<br>(%242)* | 692,17- 1505,07 |
| **Calcium (mg/d)** | | | 418,65±314,52<br>(%73)* | 268,13- 680,90 |
| **Potassium (mg/d)** | | | 1572,60±314,52<br>(%40)* | 1046,55- 2415,70 |
| **Magnesium (mg/d)** | | | 194,92±37,84<br>(%75)* | 139,90- 262,73 |
| **CBPAQ Outcome expectations** | | | 4,41±0,50 | 3,60-5,30 |
| **CBPAQ Self-regulation** | | | 3,80±0,73 | 2,80-5,00 |
| **CBPAQ Personal Barriers** | | | 2,72±1,04 | 1,00±5,00 |

|  |  |  |
| --- | --- | --- |
| **Total CBPAQ Score** | 5,49±1,93 | 2,40-8,80 |
| **WEMWBS-SF Score** | 26,24±6,56 | 12,00-35,00 |

PRAL: dietary acid load. <15 low prAL diet, >15 high PRAL diet. USG: Urine specific gravity. <1,020; euhydration, 1,020-1,030; hypohydration. BMI: body mass index. <18,5; underweight 18,5-24,9; normal weight, 25-29,9; overweight. CBPAQ: Cognitive Behavioral Physical Activity Questionnaire. WEMWBS-SF: Warwick-Edinburgh Mental Well-being Scale Short Form.
\*The ratio of average daily nutrient intake to RDA is included

The participants’ average pre-exercise weight was 57.49 ± 5.64 kg, while their mean post-exercise weight was 58.05 ± 5.68 kg. After the 12-week exercise program, a statistically significant difference was observed in the participants’ performance on the sit-reach test and left-hand grip strength (p < 0.05). However, the increase in right-hand grip strength was not statistically significant, despite the improvement (Table 2).

**Table 2.**
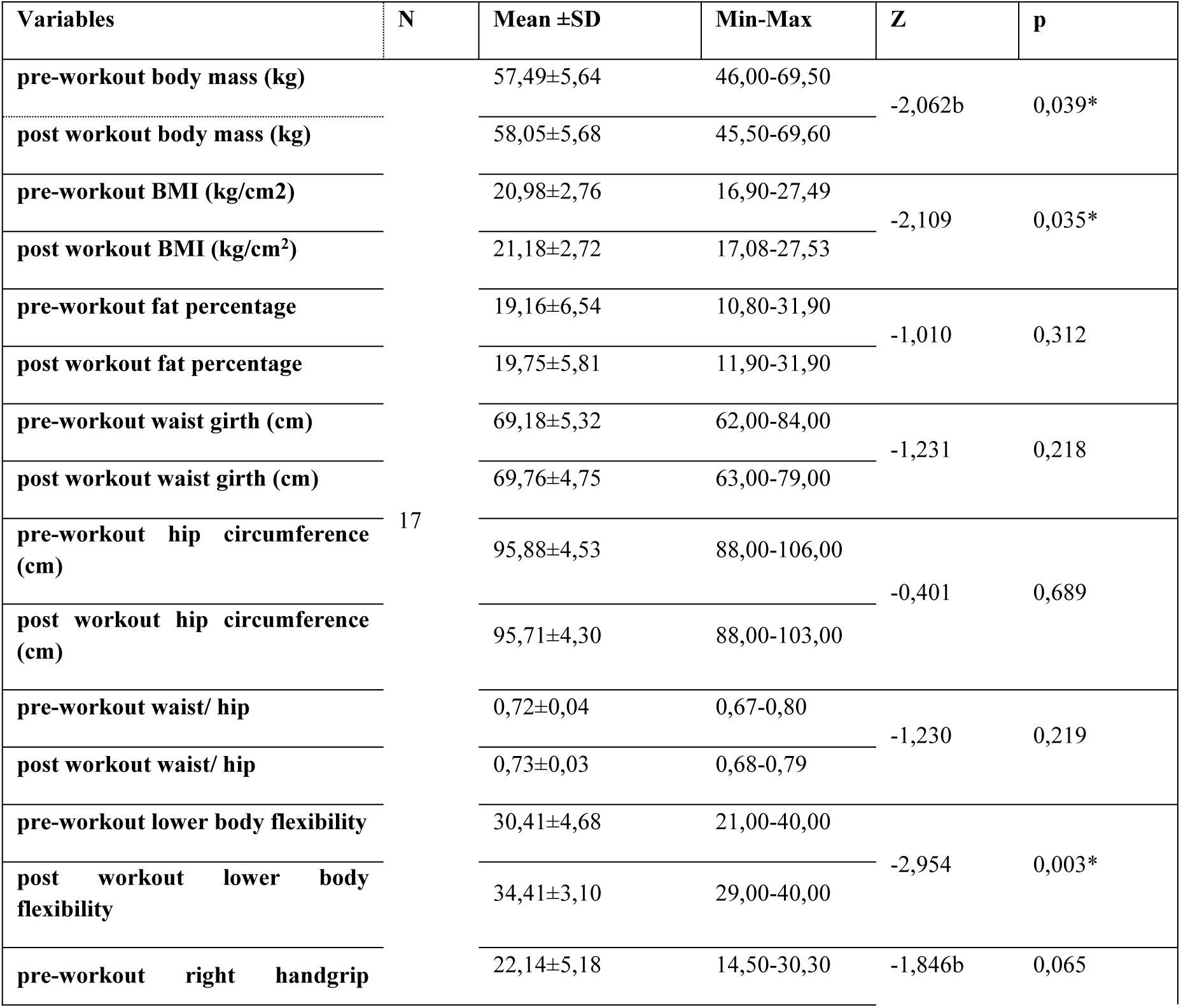
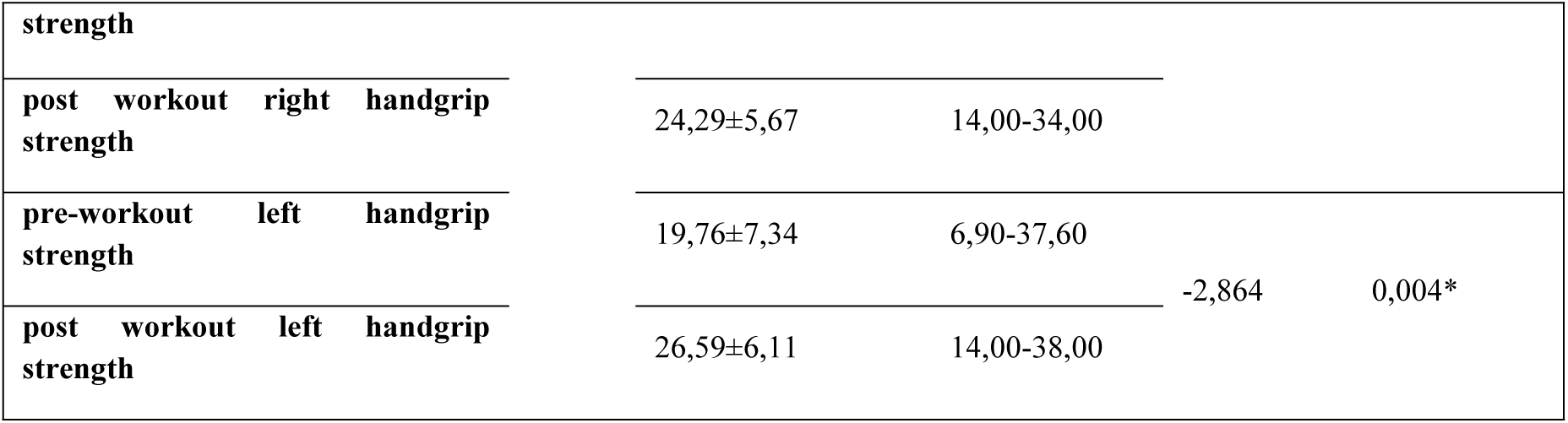
Comparison of Preliminary and Posttest Measurements for Participants’ Anthropometric Measurements and Body Composition Assessment.

Although 13 participants experienced an increase in weight and BMI, an increase in body fat percentage was observed in 9 participants, and the waist-to-hip ratio increased in 8 participants. At the end of the 12-week exercise program, a positive change in the sit-and-reach test was noted for 14 participants. When examining hand grip strength, it was found that 12 participants showed an increase in right-hand grip strength, while 14 participants exhibited an improvement in left-hand grip strength (Table 3).

**Table 3.** Comparison of Preliminary and Posttest Measurements for Participants’ Anthropometric Measurements and Body Composition Assessment.

| Variables |  | n | Rank Average | Rank Total | Z | p |
| --- | --- | --- | --- | --- | --- | --- |
| Post and Pre-workout weight (kg) | Negative Ranks | 4 | 8,25 | 33,00 | -2,062b | 0,039* |
|  | Positive Ranks | 13 | 9,23 | 120,00 |  |  |
|  | Ties | 0 |  |  |  |  |
| Post and Pre-workout BMI (kg/cm2) | Negative Ranks | 4 | 8,00 | 32,00 | -2,109 | 0,035* |
|  | Positive Ranks | 13 | 9,31 | 121,00 |  |  |
|  | Ties | 0 |  |  |  |  |
| Post and Pre- workout fat percentage | Negative Ranks | 7 | 6,93 | 48,50 | -1,010 | 0,312 |
|  | Positive Ranks | 9 | 9,72 | 87,50 |  |  |
|  | Ties | 1 |  |  |  |  |
| Post and Pre- workout waist girth (cm) | Negative Ranks | 5 | 7,70 | 38,50 | -1,231 | 0,218 |
|  | Positive Ranks | 10 | 8,15 | 81,50 |  |  |
|  | Ties | 2 |  |  |  |  |
| Post and Pre- workout hip circumference (cm) | Negative Ranks | 7 | 6,29 | 44,00 | -0,401 | 0,689 |
|  | Positive Ranks | 5 | 6,80 | 34,00 |  |  |
|  | Ties | 5 |  |  |  |  |
| **Post and Pre- workout waist/hip** | Negative Ranks | 5 | 5,60 | 28,00 | -1,230 | 0,219 |
|  | Positive Ranks | 8 | 7,88 | 63,00 |  |  |
|  | Ties | 4 |  |  |  |  |
| **Post and Pre- workout lower body flexibility** | Negative Ranks | 2 | 5,50 | 11,00 | -2,954 | 0,003* |
|  | Positive Ranks | 14 | 8,93 | 125,00 |  |  |
|  | Ties | 1 |  |  |  |  |
| **Post and Pre- workout right handgrip strength** | Negative Ranks | 5 | 7,50 | 37,50 | -1,846b | 0,065 |
|  | Positive Ranks | 12 | 9,63 | 115,50 |  |  |
|  | Ties | 0 |  |  |  |  |
| **Post and Pre- workout left handgrip strength** | Negative Ranks | 3 | 5,33 | 16,00 | -2,864 | 0,004* |
|  | Positive Ranks | 14 | 9,79 | 137,00 |  |  |
|  | Ties | 0 |  |  |  |  |

According to the data presented in Table 4, no significant difference was observed between dietary acid load, urine specific gravity, and urine pH, which are sub-dimensions of mental well-being and cognitive-behavioral physical activity, such as outcome expectations, self-regulation, and personal disabilities, in individuals who participated in Pilates exercises (p > 0.05).

**Table 4.**
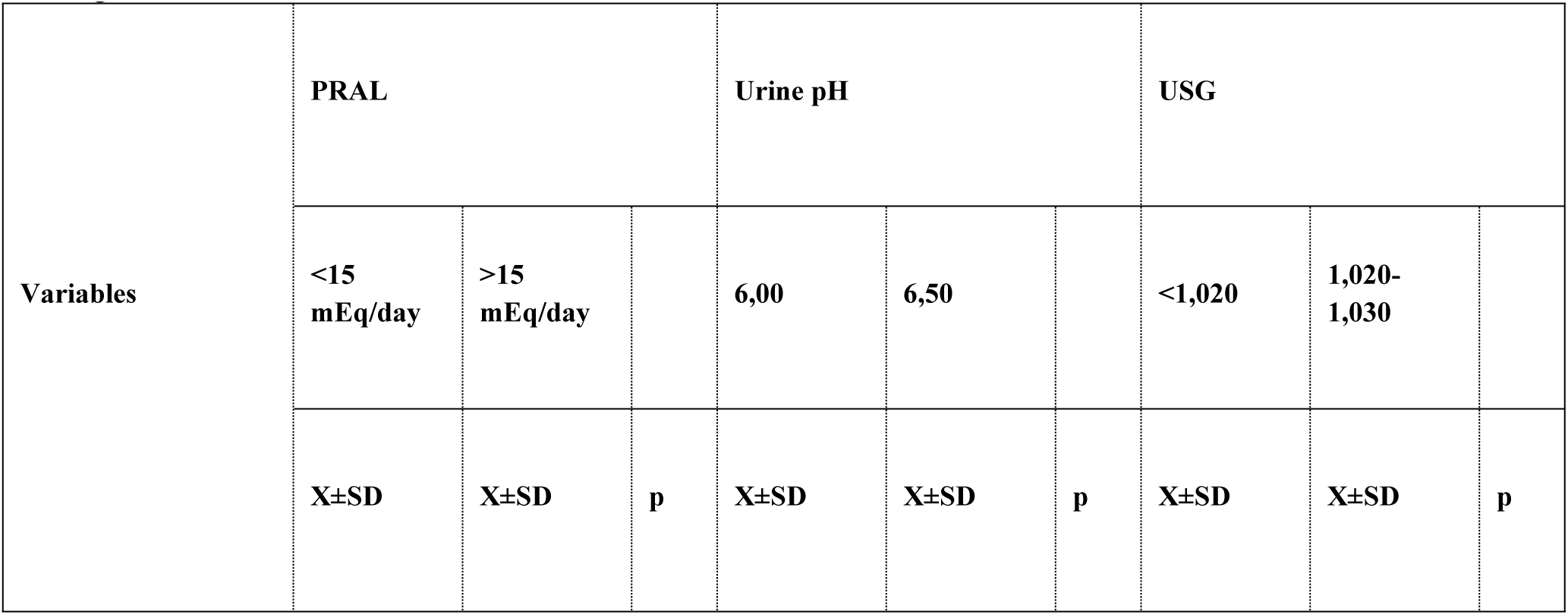
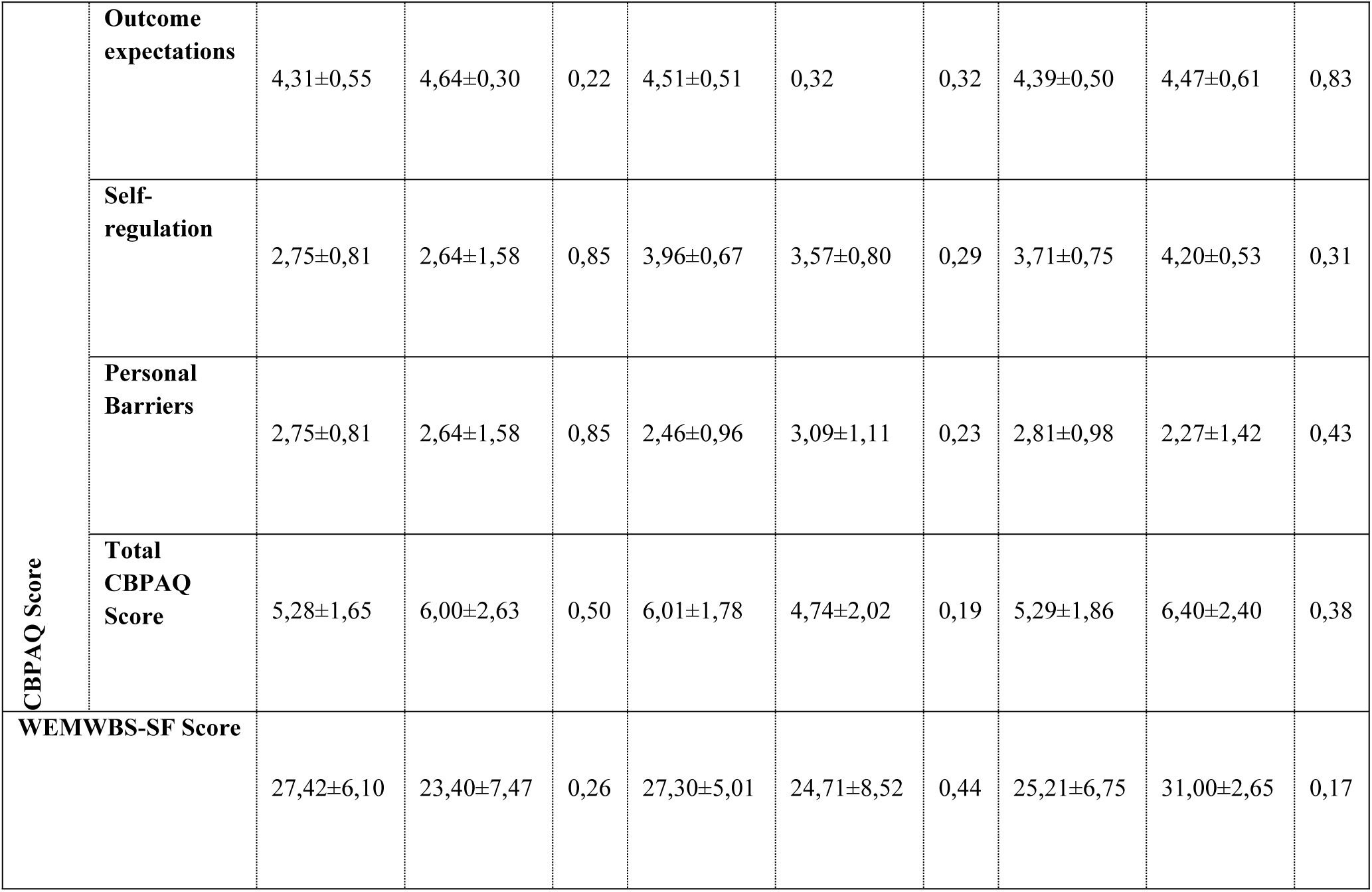
Comparison of Dietary Acid Load Variables with Physical Activity and Mental Well-Being Scores.

| Variables | PRAL |  |  | Urine pH |  |  | USG |  |  |
| --- | --- | --- | --- | --- | --- | --- | --- | --- | --- |
|  | <15<br>mEq/day | >15<br>mEq/day |  | 6,00 | 6,50 |  | <1,020 | 1,020-1,030 |  |
|  | X±SD | X±SD | p | X±SD | X±SD | p | X±SD | X±SD | p |

|  |  |  |  |  |  |  |  |  |  |  |
| --- | --- | --- | --- | --- | --- | --- | --- | --- | --- | --- |
| **CBPAQ Score** | **Outcome expectations** | 4,31±0,55 | 4,64±0,30 | 0,22 | 4,51±0,51 | 0,32 | 0,32 | 4,39±0,50 | 4,47±0,61 | 0,83 |
|  | **Self-regulation** | 2,75±0,81 | 2,64±1,58 | 0,85 | 3,96±0,67 | 3,57±0,80 | 0,29 | 3,71±0,75 | 4,20±0,53 | 0,31 |
|  | **Personal Barriers** | 2,75±0,81 | 2,64±1,58 | 0,85 | 2,46±0,96 | 3,09±1,11 | 0,23 | 2,81±0,98 | 2,27±1,42 | 0,43 |
|  | **Total CBPAQ Score** | 5,28±1,65 | 6,00±2,63 | 0,50 | 6,01±1,78 | 4,74±2,02 | 0,19 | 5,29±1,86 | 6,40±2,40 | 0,38 |
| **WEMWBS-SF Score** |  | 27,42±6,10 | 23,40±7,47 | 0,26 | 27,30±5,01 | 24,71±8,52 | 0,44 | 25,21±6,75 | 31,00±2,65 | 0,17 |

## 4. Discussion

The intake of alkaline and acidic foods leads to the formation of corresponding alkaline or acidic metabolites in the body. High protein consumption, which contributes to an increased dietary acid load, has been linked to a rise in urine-specific gravity, a key indicator of hydration status. While the precise relationship between dietary acid load (PRAL) and hydration remains underexplored, further research is needed to investigate potential connections, particularly regarding how protein intake may influence urine-specific gravity. Following the 12-week Pilates program, participants’ well-being was assessed through the lens of positive psychology. Previous studies consistently show that Pilates exercises not only improve physical health but also lead to significant enhancements in cognitive and psychological well-being. Many individuals who regularly practice Pilates report increased feelings of physical fitness and overall health. Given the crucial role of mental well-being in quality of life, this study evaluated factors such as mental well-being, attitudes toward physical activity, and behavioral consistency. These factors are essential in understanding the broader benefits of Pilates, particularly in fostering psychological resilience and overall well-being beyond physical fitness.

In athletes, higher protein intake is commonly consumed to support muscle mass growth. The recommended daily protein intake to stimulate muscle protein synthesis and maintain muscle mass typically ranges from 1.4 to 2.0 g/kg body weight (Jäger et. al., 2017). In our study, the total daily protein intake was measured at 62.38 ± 13.32 grams. Both protein and phosphorus intake were positively correlated with dietary acid load (PRAL). Protein intake in our study was found to be below the recommended dietary allowance (RDA), while phosphorus intake was significantly higher, reaching 242% of the RDA. A diet rich in dairy products and meat, which are high in protein and phosphorus, is associated with a higher PRAL, whereas a higher consumption of fruits and vegetables, which have alkalizing properties, tends to result in a lower PRAL (30). Since the protein intake of the athletes participating in this study was significantly lower than the recommended dietary allowance (RDA), a low PRAL was observed. The results of this study indicated higher dietary acidity compared to another study conducted on Iranian adults (31,32). When examining the survey of Lithuanian Olympic athletes, it is evident that both aerobic and anaerobic athletes follow a diet with a lower PRAL compared to the findings of this study (33). Among the other micronutrients influencing PRAL, an analysis of calcium, potassium, and magnesium intake revealed that potassium intake (40% of the RDA) was significantly lower (Table 1). Calcium and magnesium intake accounted for 73% and 75% of the RDA, respectively. It was hypothesized that the PRAL of the diet could also influence urine specific gravity and urine pH. Although 70% of the participants in this study followed a low PRAL diet, this was not reflected in urine pH. Additionally, when examining the participants’ urine specific gravity, the mean value was found to be in the range of 1.021-1.030, with a dehydration rate of USG 1.017 ± 0.006 (Table 1). This level of dehydration suggests that the participants’ fluid intake was insufficient to meet their hydration needs.

As a result of 12 weeks of Pilates training, participants experienced increases in BMI, fat percentage, and waist circumference. Statistically significant improvements were observed in the sit-reach test and left grip strength (Table 2). Analysis of the participants’ food consumption records revealed an increase in food intake during the evening hours, which contributed to weight gain. Following the 12-week training program, it was found that 14 participants showed improvements in the sit-reach test and left grip strength, while 12 participants demonstrated an increase in right grip strength (Table 3).In a study by Zaras et al. (2023), after 8 weeks of Pilates training with 20 women, no significant changes were observed in body mass or BMI. However, fat percentage and waist circumference showed a reduction. Additionally, while no significant changes were seen in the left and right hand grip strength, a notable improvement in lower body flexibility (sit-reach test) was reported (34). Another study shows lower body flexibility and trunk strength increase after 12 weeks of pilates training (35). Mohammadpour et al. (2020) investigated the relationship between dietary acid load and muscle strength. They found a significant increase in left- and right-hand and average muscle strength (36). On the other hand, the effect of Pilates training on total body fat percentage is not fully clarified. Although studies on obese participants have shown that mat-pilates can reduce body fat (37,38), a study conducted on healthy individuals showed that pilates may not be as effective in reducing body fat percentage (39).

The study examined the individuals’ cognitive behavioral physical activity levels, and the CBPAQ Score was 5.49±1.93. When the CBPAQ Score was examined in sub-dimensions, no significant difference was found between PRAL, urine specific gravity, and urine pH. When the study conducted by Asan et al. (2021) is examined, it is seen that all sub-dimensions and CBPAQ scores of individual athletes are lower than the study participants (40). When the study conducted by Sermenli Aydın and Keklicek (2020) on university students and the study conducted by Graded et al. (2024) examined their age groups, it was seen that the participants included in this research had higher scores in all dimensions except for the personal disabilities sub-dimension (41). As a result, it was determined that the individuals participating in this study had a cognitive perception of physical activity. In addition, WEMWBS-SF Score: There was no significant difference with PRAL, urine specific gravity, and urine pH. When the studies of Duman et al. (2020) and Tekkurşun Demir (2018) are examined, it is seen that the mental well-being scores are higher than this study finding (42,43). However, individuals gain the habit of healthy eating and regular exercise. Studies prove that it has an essential place in terms of the formation and improvement of mental health (44). In this context, it is thought that the mental well-being scores of the participants will increase even more if they make their nutrition cleaner and pay attention to their eating habits and regular exercise.

## 5. Conclusion

Contrary to our initial hypothesis, no significant associations were found between dietary acid load (PRAL) and urine specific gravity (USG). However, the results of the 12-week Pilates intervention revealed notable and transformative changes in participants’ body composition, including substantial increases in BMI, body fat percentage, and waist circumference. Significant improvements were observed in the sit-reach flexibility test and left-hand grip strength, highlighting the profound physical adaptations resulting from the Pilates training. Notably, participants also displayed a heightened cognitive perception of physical activity, suggesting that Pilates not only influences physical health but also fosters a deeper psychological connection with exercise.

Despite these encouraging findings, several limitations must be acknowledged. These include the relatively small sample size, the non-elite status of the participants, the uncontrolled nature of dietary intake, and the suboptimal sensitivity of the urine pH measurement (with a 0.5-unit sensitivity, rather than the desired 0.1-unit precision). Nevertheless, this study offers valuable insights and lays the groundwork for future research. Future studies should include larger and more diverse cohorts—incorporating male athletes and participants from various competitive levels—to further explore the complex relationship between dietary acid load and physical performance across different sports. Additionally, future training programs should be carefully tailored to the individual needs of athletes, with a focus on optimizing body composition and establishing sustainable, sport-specific nutrition habits that enhance overall performance.

## Author contributions

Conceptualization: EA, HEY, DGK

Formal analysis: HEY

Methodology: HEY, DGK

Project administration: EA, HEY, DGK

Resources: HEY

Software: EA, HEY, DGK

Validation: DGK

Visualization: EA

Writing – original draft: EA, HEY, DGK

Writing – review & editing: HEY

## Data availability

The datasets analyzed in the current study are available from the corresponding author on reasonable request.

## Acknowledgements

Authors have no acknowledgments to declare.

## Ethics Statement

In order to conduct the research, the necessary scientific research permissions were obtained from the Burdur Mehmet Akif University University Non-Interventional Clinical Research Ethics Committee (Non-Interventional Clinical Research Ethics Committee Decision, Meeting Date: 03.12.2024, Meeting No: 2024/12, Decision No: GO 2024/639).The procedures followed in this study adhered to the ethical guidelines set forth by the institutional review boards and the 1964 Helsinki Declaration, along with its subsequent amendments, or equivalent ethical standards.

## Consent to Participate

All participants were informed about the purpose of the study and voluntarily agreed to participate. Written informed consent was obtained from each participant prior to their inclusion in the study.

